# Association of Vaccine Hesitancy with Demographics, and Mental Health – United States Household Pulse Survey Study

**DOI:** 10.1101/2022.04.13.22273843

**Authors:** Arinjita Bhattacharyya, Shikshita Singh, Swarna Sakshi, Anand Seth, Shesh N Rai

## Abstract

**Background:** The world is witnessing a pandemic caused by the novel coronavirus named Covid-19 by WHO that has claimed millions of lives since its advent in December 2019. Several vaccine candidates and treatments have emerged to mitigate the effect of virus, along with came an increased confusion, mistrust on their development, emergency authorization and approval process. Increased job losses, jump in divorce rate, and the generic nature of staying home has also led to various mental health issues.

**Methods:** We analyzed two publicly available datasets to better understand vaccine hesitancy. The first dataset was extracted from ICPSR Covid-19 database (https://doi.org/10.3886/E130422V1).[1].This cross-sectional survey was conducted to assess the prevalence of vaccine hesitancy in the US, India, and China. The second dataset was obtained from the United States Census Bureau’s Household Pulse Survey (HPS) Phase 3.2.

For the ICPSR dataset, proportions and summary statistics are reported to give an overview of the global picture of vaccine hesitancy. The HPS dataset was analyzed using multinomial and binary logistic regression. Chi-square test of independence and exploratory data analysis supplemented provided insight into the casual factors involved in vaccine hesitancy.

**Results:** *ICPSR Global Data:* For India, 1761 participants completed the survey as of November, 2020 of which 90.2% indicated acceptance of a Covid-19 vaccine. 66.4% are parents of 18 years old or younger, and 79.0% respondent has a parent 50 years or older. Vaccine acceptance rate was 99.8% among 928 out of 1761 participants who had a child. 1392 participants either had a parent or child of which 83.4% will encourage their parents and 90.5% will encourage their children to get the covid-19 vaccine. In this Indian survey, 16.2% identified as belonging to the rural population of which 51.2% showed vaccine hesitancy. A binary logistic regression model with vaccine hesitancy as a dichotomous variable showed that rural population had an odds ratio (OR) of 3.45 (p-value<0.05). Income seems to influence vaccine hesitancy, with income level of (7501-15,000 Indian Rupees (INR)/month) having an OR of 1.41 as compared to other income groups. In the US, 1768 individuals participated in the survey from August-November 2020. 67.3% respondents indicated the will to accept the vaccine. 1129 of them either had a parent or a child, of which 67.6% will take the vaccine; 66% will encourage their parents and 83% will encourage their children for taking the vaccination. 40.3% responded as vaccine hesitant, 31% identified as staying in rural areas, of which 52.5% are vaccine hesitant. In the binary logistic regression analysis, race, past flu shot history, rural living, income turned out to be significant. White race had OR >1 as compared to other races, low-income group (US dollar $2000-4999/month) had an OR of 1.03. In China, there were 1727 participants, of which 1551(90.0%) indicated that they will accept a vaccine. 90.1% of them who had either a parent or child will accept vaccine, 80.4% will influence parents, and 83.4% will encourage children to get vaccination needle in the arm. 30% had vaccine hesitancy. 262 belonged to the rural population, of which 34.8% are vaccine hesitant. Income and Northern region (OR = 3.17) were significant in saying “yes” to a vaccine. High income groups were least resistant (OR=0.96) as compared to other groups.

*HPS USA data:* Data used in this study was collected from United States Census Bureau’s Household Pulse Survey (HPS) Phase 3.2 Weeks 34-39, which covers data collected from July 21, 2021, to October 11, 2021. The HPS data helped to understand the effect of several demographic and psychological, and health-related factors upon which responses were provided, thus helping to understand the social and economic effects during the COVID-19 pandemic.

**Conclusion:** Among the three countries, it appears based on this survey that US has the highest rate of vaccine hesitancy. may contribute towards this result gender, education, religious beliefs, disbelief in science, government which remains unexplored due to data limitation.

## Introduction

Vaccine hesitancy has posed challenge for health care providers and public health officials for years. In the light of the COVID-19 pandemic, vaccine hesitancy is more relevant than ever and has become a global issue. The SAGE Working Group on Vaccine Hesitancy (WG) defines vaccine hesitancy as a “delay in acceptance or refusal of vaccination despite availability of vaccination services” [2]. There are several conceptual models for the grouping of vaccine hesitancy, WG incorporated the “3 Cs” model into their definition of vaccine hesitancy. The “3 Cs” model consists of (1) confidence, (2) complacency, and (3) convenience. Confidence represents the faith of the recipient in the effectiveness and safety of the vaccine and in the delivering system. Complacency occurs when the perceived risk of the disease is low in view of the recipient; other factors can also contribute to complacency, such as initial success of the vaccine program. Convenience relates to the availability and accessibility of the vaccine of the desired vaccine [2].

Vaccine hesitancy is not unique to the SARS-CoV-2 vaccine. Despite high childhood vaccination rates in developed countries, recent outbreaks of vaccine preventable diseases, such as measles and mumps, have demonstrated the existence of clusters of unvaccinated populations (Dube). A national survey of childhood vaccines and the influenza vaccines in the Unites States found that 1 in 15 parents were hesitant about childhood vaccines, while the prevalence for influenza vaccine hesitancy was more than 1 in 4 parents. Additionally, the same survey found that about 1 in 4 parents believed the influenza vaccine to be effective. One in 8 parents were also concerned with the side effects of the influenza and routine childhood vaccines (Kempe). Prevailing hesitancy towards vaccines in the United States begs the question: What are the specific factors that contribute to vaccine hesitancy?

It was attempted to classify the barriers to the influenza vaccine uptake into its micro- and macro-levels[3]. The micro-level barriers are generally psychological and physical that can be related to theories of health decision making and behavior. The authors identified 258 micro-level barriers. These barriers were subsequently grouped in the following categories: utility, risk perception, social benefit, subjective norm, perceived behavioral control, attitude, past behavior, experience, knowledge, and unhealthy lifestyles. Specifically, our focus will be on respondents’ attitude, sociodemographic factors, and their risk perception. Additionally, we will break down the COVID-19 vaccine hesitancy rates among the different regions of the United States.

The contributing factors for COVID-19 vaccine hesitancy can vary. Social media organization [4] vaccine characteristics [5] political affiliations [6], education level[7], employment, risk of infection [8]distrust of the COVID-19 vaccine [9], and general vaccine avoidance [10] have been associated with COVID-19 vaccine hesitancy. Past studies looking into COVID-19 vaccine hesitancy have used a large-n cross-country regression framework, survey with choice-based conjoint analysis, and regression analyses have been used to analyze data in previous studies. Our study aims to provide an estimation of the COVID-19 vaccine hesitancy rate while using the U.S. Census Bureau’s Household Pulse Survey’s (HPS) hesitancy responses as the dependent variables for the multinomial and binary logistic regression models.

### Literature Study

“Survey studies on COVID-19 vaccine acceptance rates were found from 33 different countries. Among adults representing the general public, the highest COVID-19 vaccine acceptance rates were found in Ecuador (97.0%), Malaysia (94.3%), Indonesia (93.3%) and China (91.3%). However, the lowest COVID-19 vaccine acceptance rates were found in Kuwait (23.6%), Jordan (28.4%), Italy (53.7), Russia (54.9%), Poland (56.3%), US (56.9%), and France (58.9%)”[11].

A recent systematic review on the impact on mental health during Covid-19 reported that there have been relatively high rates of symptoms of anxiety (6.3% to 50.9%), depression (14.6% to 48.3%), post-traumatic stress disorder (7.0% to 53.8%), psychological distress (34.4% to 38.0%), and stress (8.1% to 81.9%) among global population of China, Spain, Italy, Iran, the US, Turkey, Nepal, and Denmark [12]. Other studies also found high rates of negative mental health outcomes in the Italian general population within 3 weeks of lockdown[13]. Studies have also shown a positive association between the probability of contracting COVID-19 and anxiety within the younger age group, while older people were better in this perspective and did not show concerns regarding mental health [14]. Experts have pointed out the need and attention to covid-19 related impacts on mental health, stress, anxiety, and human psychology. Providing psychological first aid is an essential care component for populations that have been victims of emergencies and disasters, before, during and after the event[15].

Another study conducted in China finds that the high prevalence of mental health problems, was positively associated with frequent social media exposure during the COVID-19 outbreak that requires further attention from health and government officials [16]. The anxiety levels increased from 18.1% before the pandemic to 25.3% within four months after the pandemic began; and the prevalence of moderate-severe depression increased from 21.5% to 31.7%.

### Data Description

Here we analyze two cohorts 1) ICPSR Covid-19 database which focuses on the global vaccine hesitancy data 2) Household Pulse Survey data that concentrates on surveys of identified households in the USA. A detailed overview of both the datasets are addressed below:

#### ICPSR data

The first dataset is extracted from ICPSR Covid-19 database (https://doi.org/10.3886/E130422V1) [1]The cross-sectional survey is conducted to assess the prevalence of vaccine hesitancy in the US, India, and China, due to their large sample sizes. For India, there were 1761 participants who completed the survey as of November, 2020 of which 90.2% indicated acceptance of a Covid-19 vaccine. In the US, 1768 people participated in the survey from August-November 2020. Of the people participated, 67.3% indicated that they will accept the vaccine. 1129 of them either had a parent or a child, of which 67.6% will take the vaccine. In China, there were 1727 participants, of which 1551 indicated that they will accept a vaccine. 90.1% of them who had either a parent or child were willing to accept a vaccine.

#### HPS data

Data used in this study was collected from United States Census Bureau’s Household Pulse Survey [17](HPS) Phase 3.2 Weeks 34-39, which covers data collected from July 21, 2021, to October 11, 2021. We elected to use the HPS dataset because it has a number of variables that provide a measure of household experiences and social and economic effects of the COVID-19 pandemic. HPS data has several features and from a large sample across the country. Using the HPS data and using multinomial logistic regression analysis, we hope to characterize the key aspects contributing to COVID-19 vaccine hesitancy across different demographics and regions of America.

The data set contains 515,558 household samples consisting of weeks 34-39 and 202 characteristics, of which 382,908 were individual households. The dataset that was used for the analysis consisted of those who did not receive any vaccine which comprised of 43859 (11.4%) samples and 202 variables. After removing the missing values, the population reduced to 5758 samples and 202 factors. Those households responded to Question seen but category not selected (−99) and Missing/Did not report (−88) were not considered. Finally, the dataset comprised of 5758 unique participant households. Based on the previous literature on vaccine hesitancy and inspection of the datasets the following covariates were chosen to understand their relationship with vaccine hesitancy: gender, region, health insurance, income, mental health service status, marital status, mental health medicine prescribed, race, indicators of anxiety, interest in work, feeling down, worried, education, seeing, hearing, mobility, remembering impairments. Additionally, we defined two more variables one for the region (Northeast, Mid-Atlantic, Southeast, Great Lakes, Midwest, Southwest, Northwest, West, Mid-South) and the other for the political views (red, blue, red/blue) depending on the past decade of election results.

## Methods

Here we analyze two datasets on vaccine hesitancy. The first dataset is extracted from ICPSR Covid-19 database (https://doi.org/10.3886/E130422V1). The cross-sectional survey is conducted to assess the prevalence of vaccine hesitancy in the US, India, and China. The second dataset was extracted from the United States Census Bureau’s Household Pulse Survey (HPS) Phase 3.2.

For the the ICPSR dataset, we report proportions and summary statistics to give an overview of the vaccine hesitancy global picture. The HPS dataset was analyzed using multinomial and binary logistic regression. Individual Chi-square test of independence between vaccine hesitancy and health categories, and exploratory data analysis supplemented and helped in our understanding of the casual factors influencing vaccine hesitancy. RStudio and Excel were utilized for the analysis.

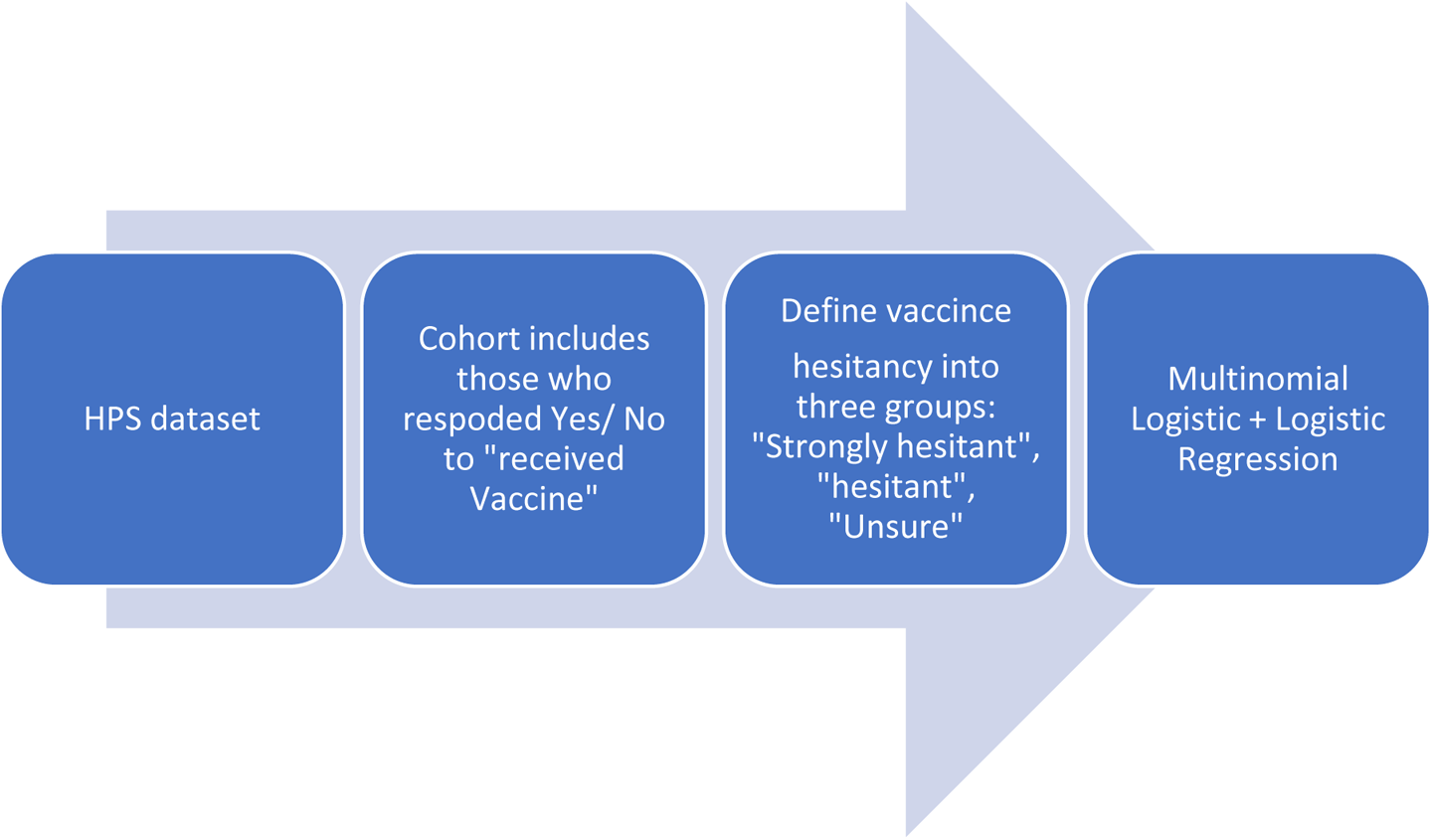

Multinomial logistic regression is used to model nominal outcome variables that have more than two categories, in which the log odds of the outcomes are modeled as a linear combination of the predictor variables (*X*_1_, … ., *X*_*p*_).

Supervise Data Mining

## Model 1

Multinomial logistic regression is used to predict categorical placement in or the probability of category membership on a dependent variable based on multiple independent variables. The independent variables can be either dichotomous (i.e., binary) or continuous (i.e., interval or ratio in scale). Multinomial logistic regression is a simple extension of binary logistic regression that allows for more than two categories of the dependent or outcome variable. It utilizes maximum likelihood estimation to estimate the coefficients. Here P (Hesitant)=p_1_= probability of falling in class “Hesitant”, P (Unsure)=p_2_= probability of falling in class “Unsure”, P (Not Hesitant) =p_3_= probability of falling in class “Not Hesitant”, p_1_+p_2_+p_3_=1

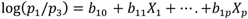

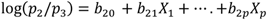

After obtaining the estimates of the coefficients (*b*_10_, *b*_11_, … ., *b*_1*p*_, *b*_20_, … . *b*_2*p*_), the odds ratios (OR) which are obtained by inverting the log odds, are used to determine the significance of the independent variables when compared between the two groups. If OR>1, and p-value<0.05, then the participants are more likely to be in the Hesitant (Unsure) group than the Not-Hesitant group.

## Model 2

Binary logistic regression is a subset of the multinomial logistic regression in which there are only two categories for the response variable. Here there are only 2 categories in the outcome variable Hesitant, and Not hesitant. The Unsure group has been divided into two other groups.

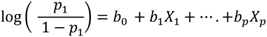

## Model 3

The dataset was divided into training and testing datasets and the multinomial logistic regression model was applied and the response were predicted on the test dataset. Confusion matrix was created to address the accuracy, sensitivity, and specificity of the model [18]

## Model 4

### Penalized Methods

In penalization methods, feature selection and efficient classifier construction are achieved simultaneously. Among the penalized techniques Least Absolute Shrinkage and Selection Operator (Lasso), Elastic-Net (EN), and Ridge Regression are widespread[19].

#### Lasso

It regularizes the regression coefficients toward zero by penalizing the regression model with the sum of coefficients as a penalty term called L1-norm. This penalty forces the coefficient estimates, with a minimum contribution to the model, to zero.

The log-likelihood of LR is shown below:

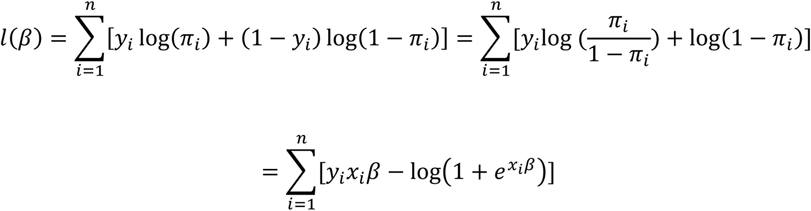

When using Lasso penalty term *λ*, the likelihood looks like this:

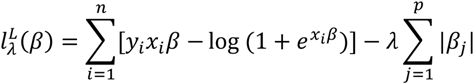

- It uses the L1 penalty, uniformly penalizing all the parameters.
- The fit is independent of multiplicative scaling.

#### Ridge

Ridge assigns the L2 penalty that is the squared magnitude of the overemphasized coefficients with λ determining the weight assigned to the penalty. The larger the value of λ, the more likely the coefficients approach zero. Unlike Lasso, the Ridge model will not shrink these coefficients to precisely zero. The likelihood is:

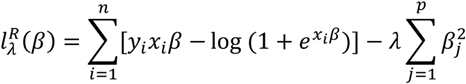

Here, β2 is the L2 penalty.

- It uses the L2 penalty, penalizing insignificant coefficients more.
- It is indifferent to multiplicative scaling.

#### Elastic - Net

EN is a convex combination of Lasso and **Ridge**, with the effectual reduction in the effect of coefficients with L2 norm and exactly setting some coefficients to zero with L1 norm. The likelihood is:

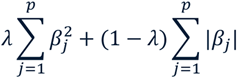

Here *λ* is the penalty as a mixture of the previous two approaches.

- It appears to be complicated with scaling.
- It can outperform Lasso on data with positively correlated variables.

## Results

### Pre-Processing

The responses towards intention of getting vaccine was categorized into the following groups: 1) Definitely get a vaccine as “Not Hesitant” 2) Probably get a vaccine and be unsure about getting a vaccine as “Unsure” 3) Probably Not get a vaccine as “Hesitant”, 4) Definitely not get a vaccine as “Strongly Hesitant”. The study is conducted to understand in-depth factors responsible for vaccine hesitancy. The sample size is adequate to infer relating to a broader population.

For the application of multinomial logistic regression analysis, we have considered Hesitant and Strongly Hesitant category in one group. The income categories were redefined as “low-income group” if the total household income is <$34, 999, “middle income group between $34,999 - $74,999, and high-income group who earned greater than $75,000. The education categories were reclassified into a) “High school” (less than high school and some high school), b) “High school graduate” (high school graduate or equivalent), c) “some college, no degree received”,” Associate/Bachelor’s Degree”, “Graduate Degree”. The response relating to depression factors such as frequency of anxiety, worry, interest, down over past 2 weeks were regrouped into three categories: “Not at all”, “Several days”, and “Always” comprises of responses for (“More than half the days”, “Nearly every day”). Other forms of impairment such as hearing, seeing, remembering, mobility was also included and recategorized as “Impaired” (Some difficulty, a lot of difficulty, cannot do at all), and “Not Impaired” (No difficulty).

Other than these chosen factors for investigation in the multinomial logistic regression model to understand their impact towards vaccine hesitancy, we also chose race (White, black, others); gender (male, female, transgender); whether the respondents holds health insurance (Yes, no); whether the respondents received mental health (MH) services (Yes, no), whether mental health medicines (psycho pharmacological drugs?) were prescribed (Yes, no); region (Midwest, northeast, south, west). People in Midwest and west regions seemed to be more vaccine hesitant than those in the northeast or south regions. Further these regions were divided by topography into eight regions as West, Northwest, Midwest, Southwest, Southeast, Mid-Atlantic, Northeast, and Great Lakes. Also, for further subset were identifies among these states, which were divided into red, red/blue or swing states, and blue states, depending on the results of presidential election over the past decade. All the missing values were omitted and the data we utilized for the analysis included 5758 respondents, and p=18.

Table 1 describes the summary statistics of baseline characteristics, among the three hesitancy groups. The Chi-square test of significance and p-values are reported. Gender, mental health services, income, race, stress indicators such as anxiety, worry, interest, down, education, marital status, state, visual impairment, mobility, and remembering (recall?) were statistically significant (p < 0.05). Figure 1 shows the counts of the vaccine hesitancy status of the households in the USA. Maximum proportion of the households were hesitant followed by Unsure about taking a vaccine.

**Table 1.**
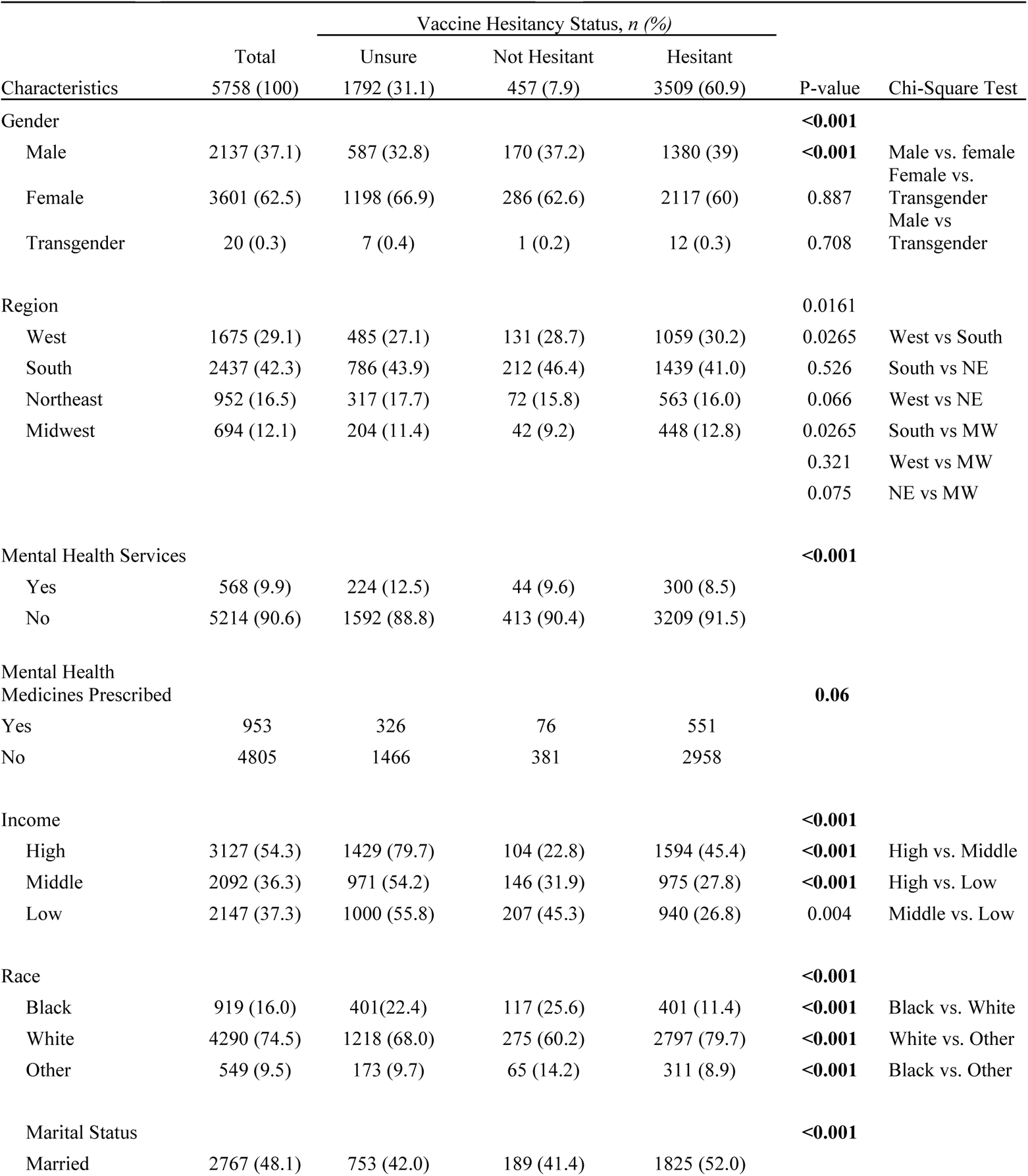
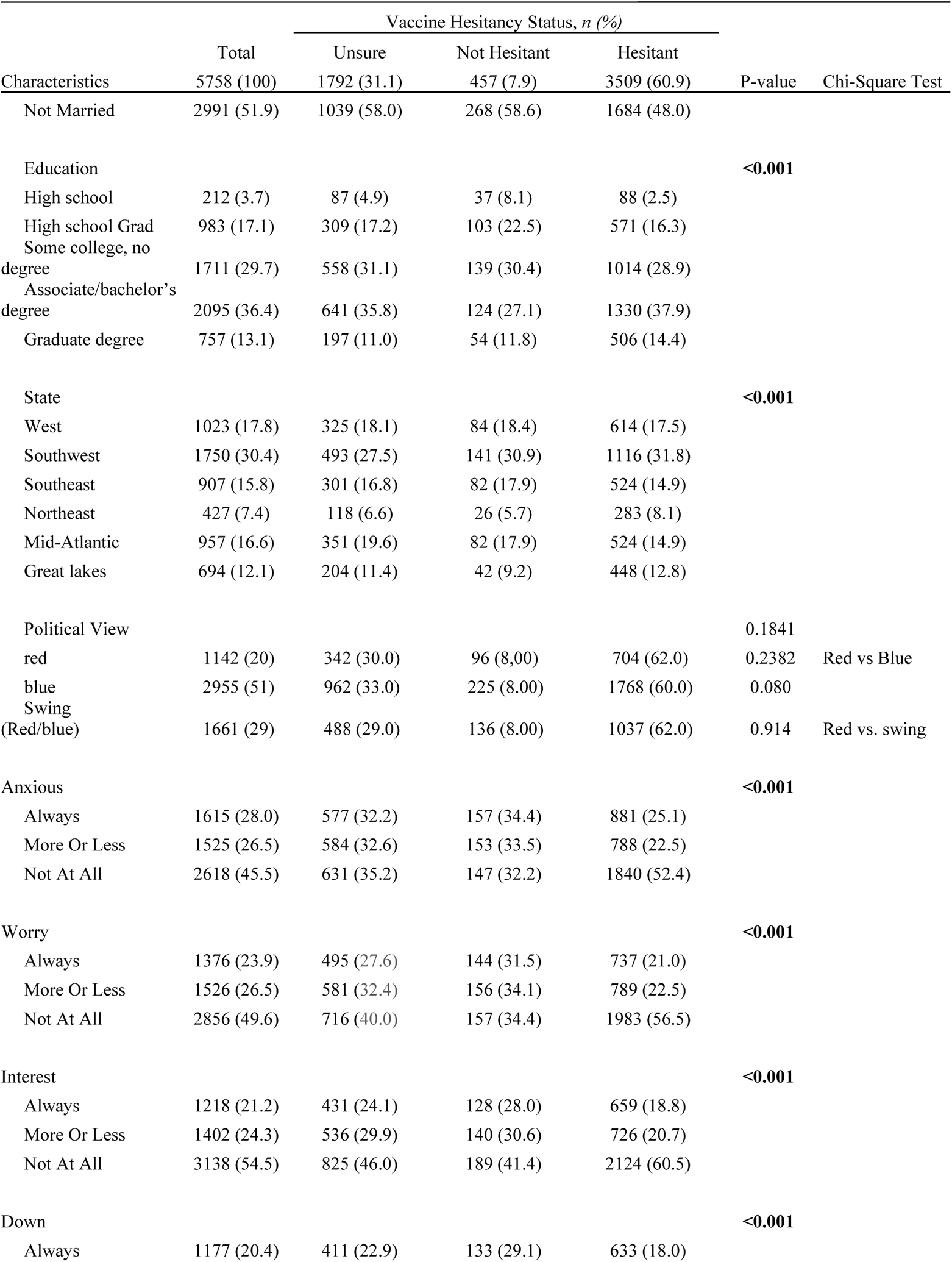
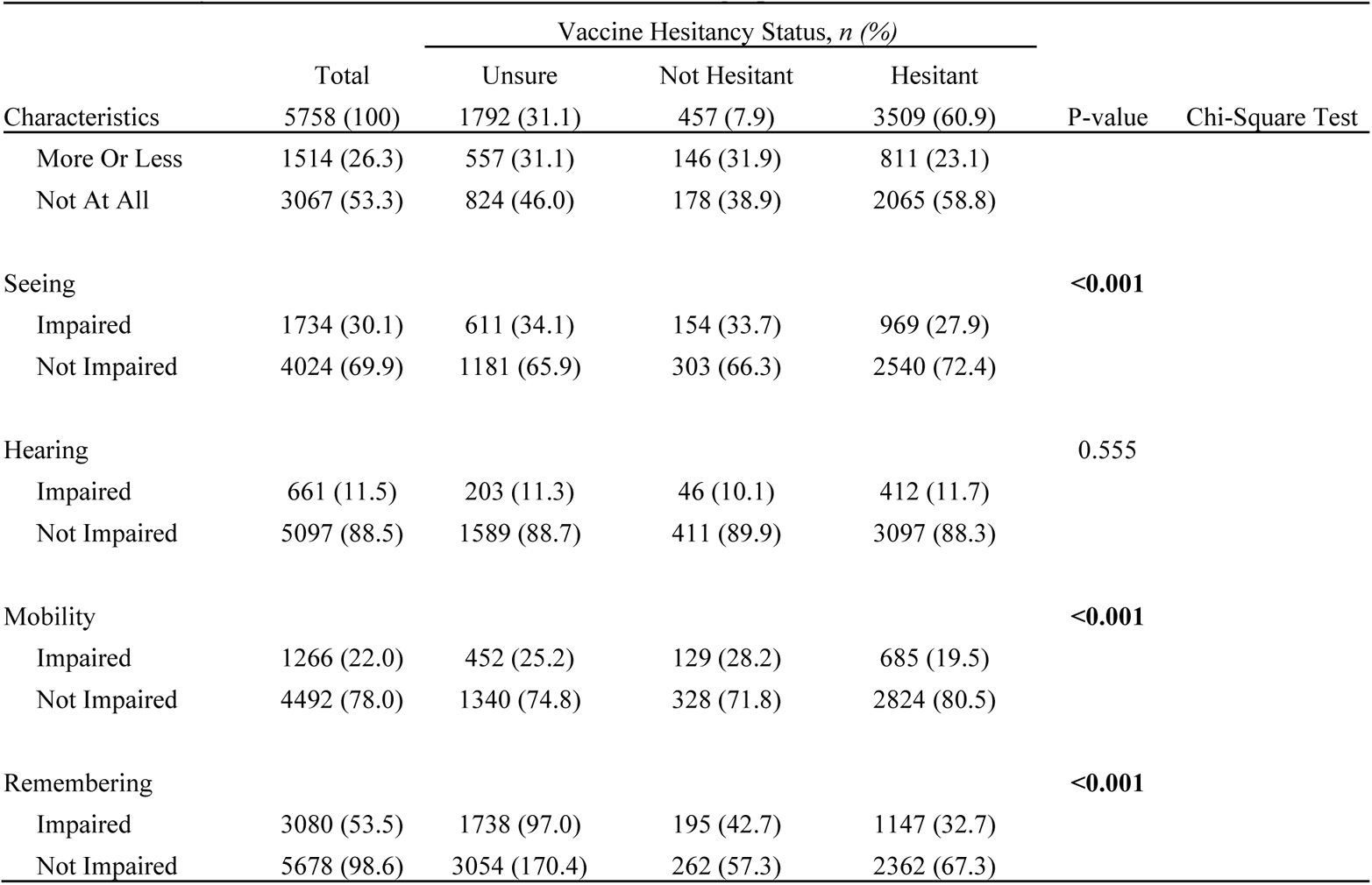
Summary Statistics Baseline Characteristics and Demographics

### Primary Results

Those who did not receive MH services had higher percentages of vaccine hesitancy (strongly hesitant+ hesitant ∼61%) than those who received MH services (strongly hesitant+ hesitant ∼56%) (Figure 1) Females receiving MH services seemed to be more vaccine hesitant than who did not receive any MH services.

Model 1: Multinomial Logistic Regression

Considering previous literatures, and other vaccine hesitancy studies, we selected the above-mentioned factors that would contribute towards understanding the outcome of interest (vaccine hesitancy). Model 1 considers the full model including all the samples and the 17 chosen variables.

We considered the variables to be significant if p <0.05 and with OR >1 compared to the reference group. Also, we created a joint p-value called a combined p-value that gathers the information from both the comparisons and is utilized for testing the overall result. The cut-off for this combined p-value is also 0. 05.

In Model 1, the multinomial logistic regression analysis is carried out, in which we have considered Hesitant and Strongly Hesitant category in one group. This is the full model. The computational algorithm converged thereby providing the estimates of the coefficients. In Model 2, the binomial logistic regression was applied with two groups. In Model 3, the dataset was split into training and testing datasets, and the prediction accuracies, sensitivities and specificities are reported. In all the models, “Not Hesitant” was considered as the reference category. Results from Model 1 multinomial logistic regression relating sociodemographic and health characteristics to the odds of belonging to three hesitancy classes (n=5758). The odds ratio estimate, 95% CI, and the P-values corresponding to the Wald test are reported. Reference categories are in parenthesis in table 2.

**Table 2:**
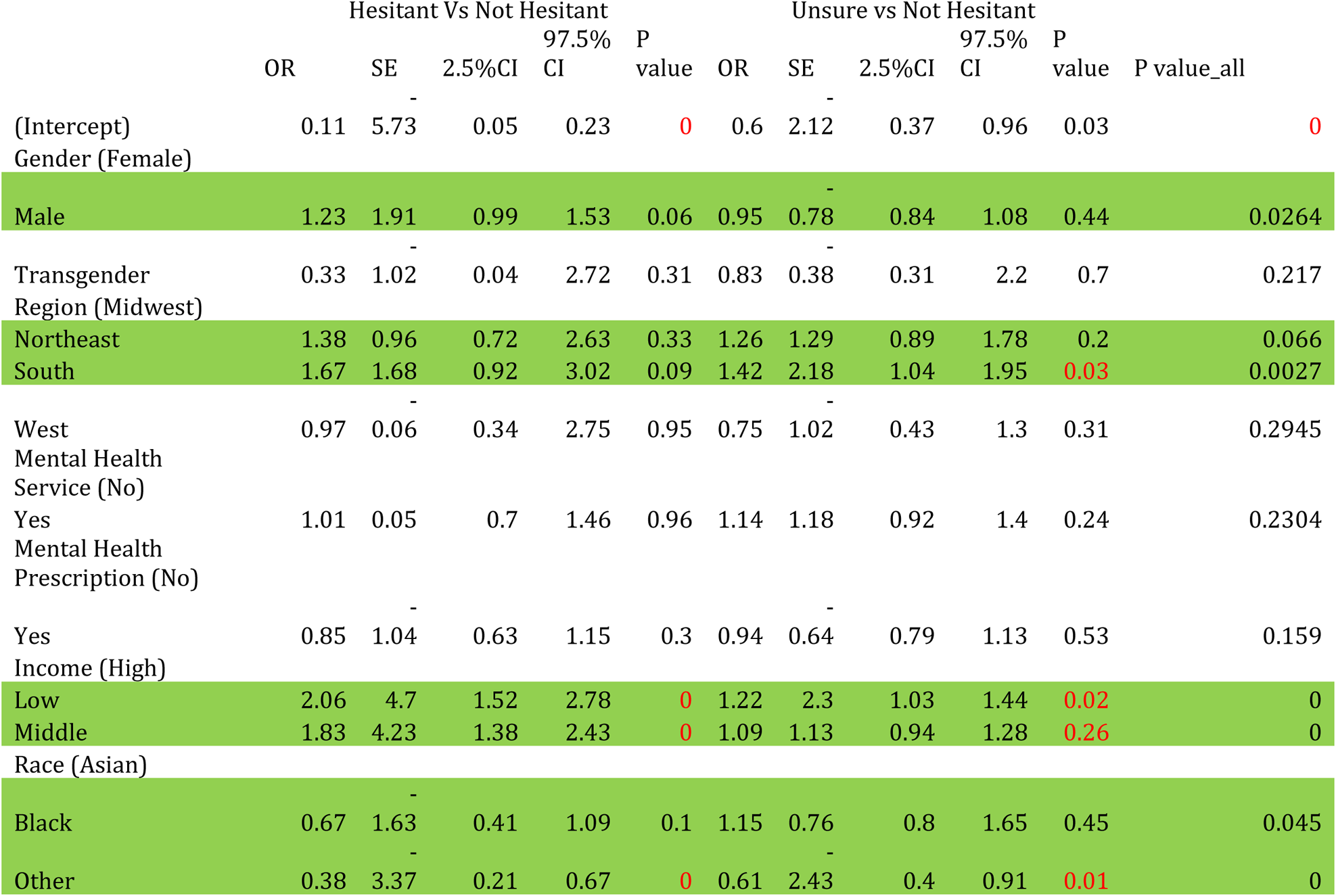
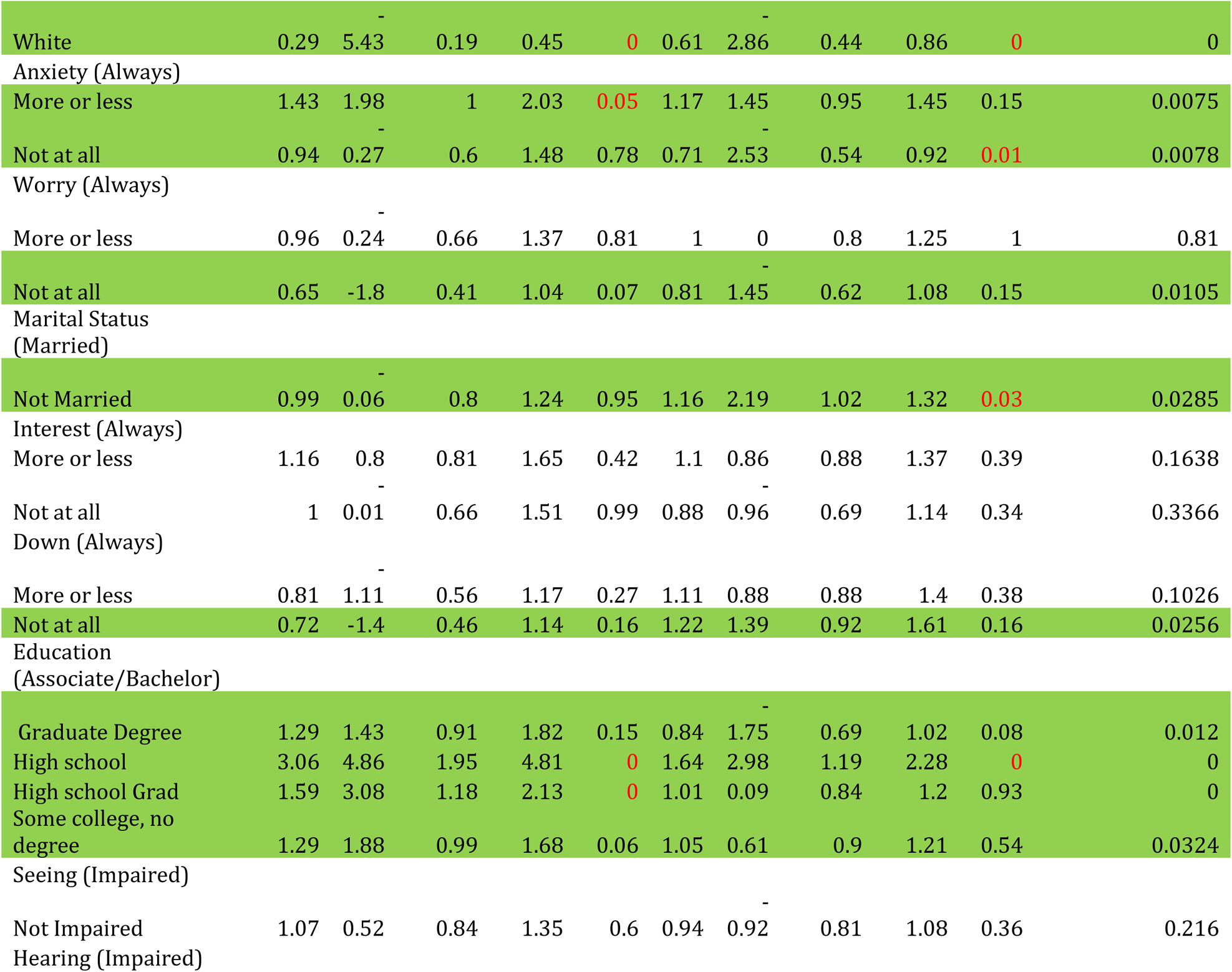
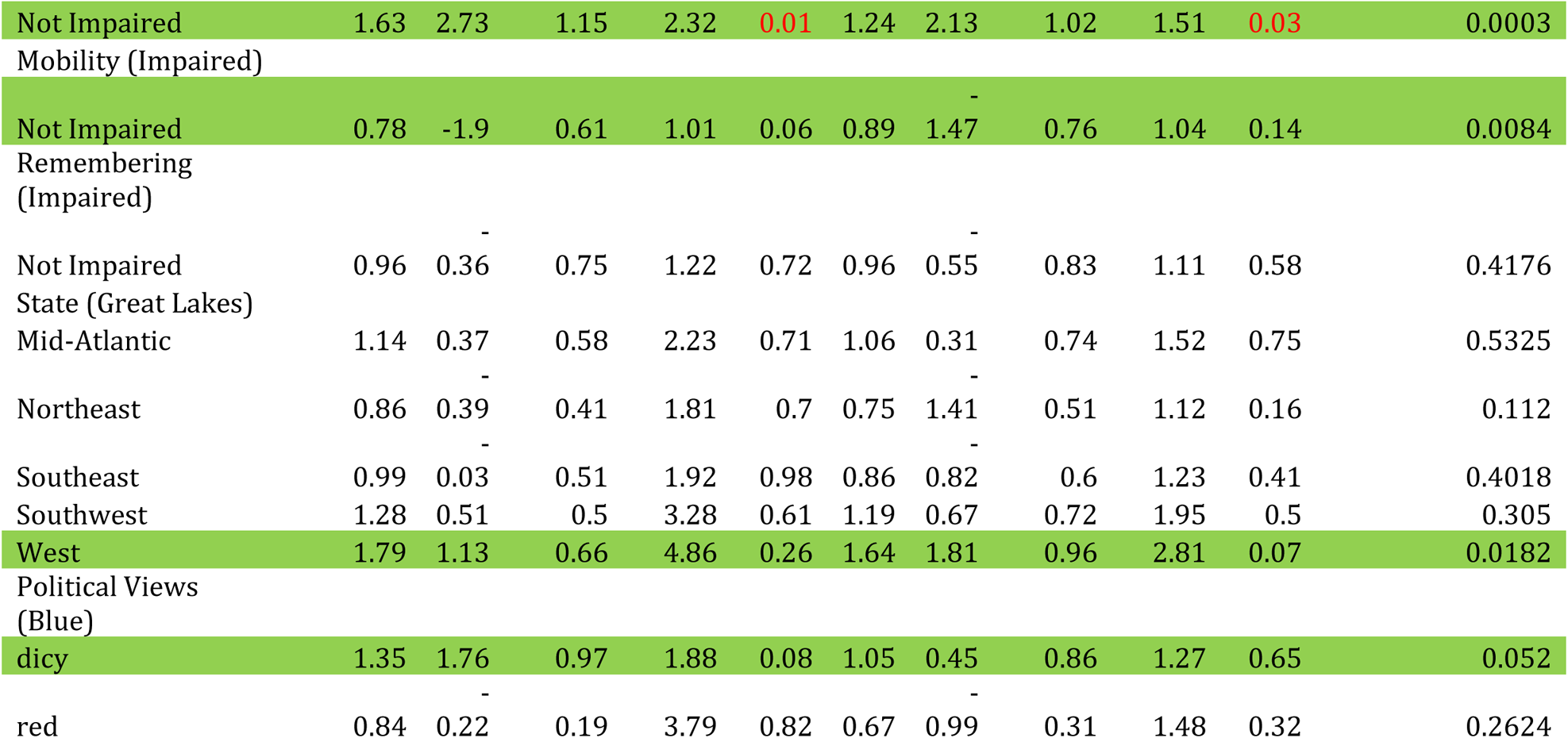
Multinomial Logistic Regression Results

Males were more likely to be hesitant or unsure towards vaccine than the females. People belonging to the northeast and south regions are more vaccine hesitant /unsure than those in the Mid-west.

Low- and middle-income groups of people were likely to be hesitant or unsure about vaccines than high income groups. Unmarried people were vaccine pro than married people. Asians are more likely to be in the not-hesitant groups than other races. Those who are not at all down or worry are more likely to be in the Not hesitant group than in the hesitant group. High school students, those having some education, no degree or graduate degree were more likely to be in the hesitant/unsure group than in the not hesitant group. Those with impaired mobility and impaired hearing belonged to the not hesitant group. Respondents from Western states were more leaning towards vaccine hesitancy than from other states. Respondents from swing states were more towards vaccine hesitancy or being unsure than non-hesitancy.

Model 2 Binomial logistic regression

In model 2, we segregated the unsure group from Hesitant and Non-Hesitant groups, thus having two outcomes, for which a binomial logistic regression model is best suited. Results from Model 2 binomial logistic regression relating sociodemographic and health characteristics to the odds of belonging to three hesitancy classes (n=5758). The odds ratio estimate, 95% CI, and the P-values corresponding to the Wald test are reported. Reference categories are in parenthesis

Model 2 had similar results with those belonging to male group, from west region, those who belonged to black, other, or white race groups, low- and middle-income groups, having high school, high school grad, some college, no degree, belonging to red states were more likely to belong to the vaccine hesitant group than the not hesitant group. People of the western region, having impaired mobility were more likely to belong to the not-hesitant group than in the hesitant group.

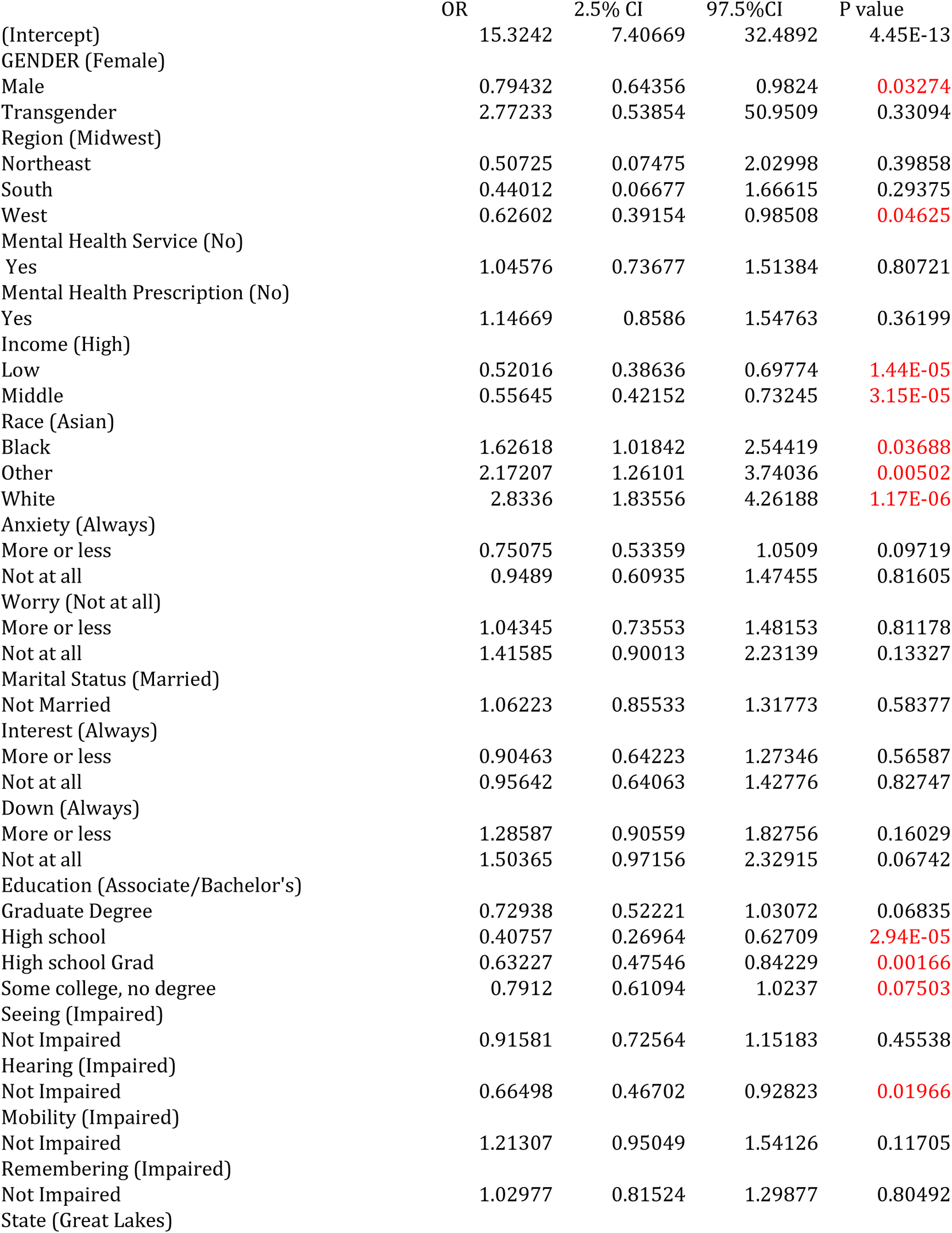
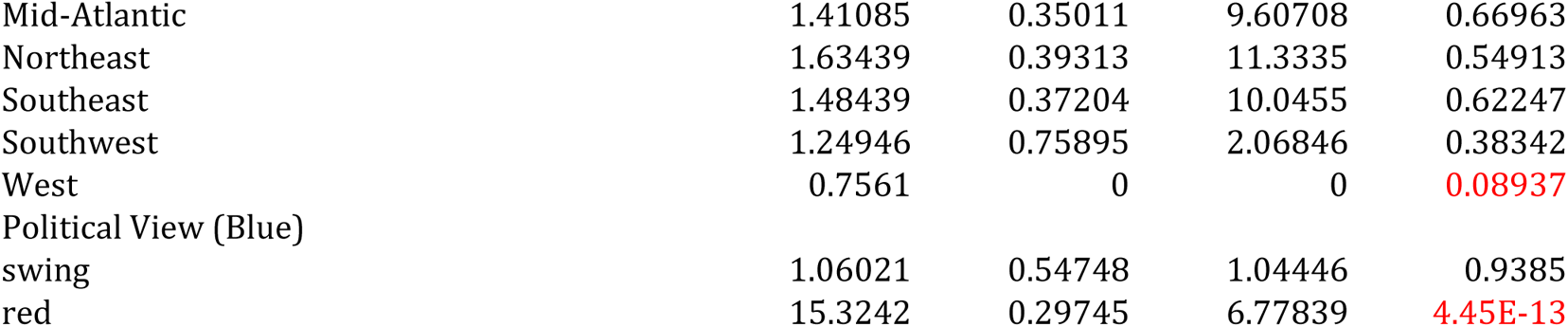

|  | OR | 2.5% CI | 97.5%CI | P value |
| --- | --- | --- | --- | --- |
| (Intercept) | 15.3242 | 7.40669 | 32.4892 | 4.45E-13 |
| GENDER (Female) |  |  |  |  |
| Male | 0.79432 | 0.64356 | 0.9824 | 0.03274 |
| Transgender | 2.77233 | 0.53854 | 50.9509 | 0.33094 |
| Region (Midwest) |  |  |  |  |
| Northeast | 0.50725 | 0.07475 | 2.02998 | 0.39858 |
| South | 0.44012 | 0.06677 | 1.66615 | 0.29375 |
| West | 0.62602 | 0.39154 | 0.98508 | 0.04625 |
| Mental Health Service (No) |  |  |  |  |
| Yes | 1.04576 | 0.73677 | 1.51384 | 0.80721 |
| Mental Health Prescription (No) |  |  |  |  |
| Yes | 1.14669 | 0.8586 | 1.54763 | 0.36199 |
| Income (High) |  |  |  |  |
| Low | 0.52016 | 0.38636 | 0.69774 | 1.44E-05 |
| Middle | 0.55645 | 0.42152 | 0.73245 | 3.15E-05 |
| Race (Asian) |  |  |  |  |
| Black | 1.62618 | 1.01842 | 2.54419 | 0.03688 |
| Other | 2.17207 | 1.26101 | 3.74036 | 0.00502 |
| White | 2.8336 | 1.83556 | 4.26188 | 1.17E-06 |
| Anxiety (Always) |  |  |  |  |
| More or less | 0.75075 | 0.53359 | 1.0509 | 0.09719 |
| Not at all | 0.9489 | 0.60935 | 1.47455 | 0.81605 |
| Worry (Not at all) |  |  |  |  |
| More or less | 1.04345 | 0.73553 | 1.48153 | 0.81178 |
| Not at all | 1.41585 | 0.90013 | 2.23139 | 0.13327 |
| Marital Status (Married) |  |  |  |  |
| Not Married | 1.06223 | 0.85533 | 1.31773 | 0.58377 |
| Interest (Always) |  |  |  |  |
| More or less | 0.90463 | 0.64223 | 1.27346 | 0.56587 |
| Not at all | 0.95642 | 0.64063 | 1.42776 | 0.82747 |
| Down (Always) |  |  |  |  |
| More or less | 1.28587 | 0.90559 | 1.82756 | 0.16029 |
| Not at all | 1.50365 | 0.97156 | 2.32915 | 0.06742 |
| Education (Associate/Bachelor's) |  |  |  |  |
| Graduate Degree | 0.72938 | 0.52221 | 1.03072 | 0.06835 |
| High school | 0.40757 | 0.26964 | 0.62709 | 2.94E-05 |
| High school Grad | 0.63227 | 0.47546 | 0.84229 | 0.00166 |
| Some college, no degree | 0.7912 | 0.61094 | 1.0237 | 0.07503 |
| Seeing (Impaired) |  |  |  |  |
| Not Impaired | 0.91581 | 0.72564 | 1.15183 | 0.45538 |
| Hearing (Impaired) |  |  |  |  |
| Not Impaired | 0.66498 | 0.46702 | 0.92823 | 0.01966 |
| Mobility (Impaired) |  |  |  |  |
| Not Impaired | 1.21307 | 0.95049 | 1.54126 | 0.11705 |
| Remembering (Impaired) |  |  |  |  |
| Not Impaired | 1.02977 | 0.81524 | 1.29877 | 0.80492 |
| State (Great Lakes) |  |  |  |  |
| Mid-Atlantic | 1.41085 | 0.35011 | 9.60708 | 0.66963 |
| Northeast | 1.63439 | 0.39313 | 11.3335 | 0.54913 |
| Southeast | 1.48439 | 0.37204 | 10.0455 | 0.62247 |
| Southwest | 1.24946 | 0.75895 | 2.06846 | 0.38342 |
| West | 0.7561 | 0 | 0 | 0.08937 |
| Political View (Blue) |  |  |  |  |
| swing | 1.06021 | 0.54748 | 1.04446 | 0.9385 |
| red | 15.3242 | 0.29745 | 6.77839 | 4.45E-13 |

Model 3: Training & Test Data

In this model, the data was partitioned having n1% of data as the training set and n2%=1-n1% as the test set. The accuracy, sensitivity, specificity, and model performance are given in Table 4. With 85% data reserved for the training set, and rest for the test set, the model has the highest accuracy of 63.5% in the training set, and 70.12% in the test set.

**Table 4:**
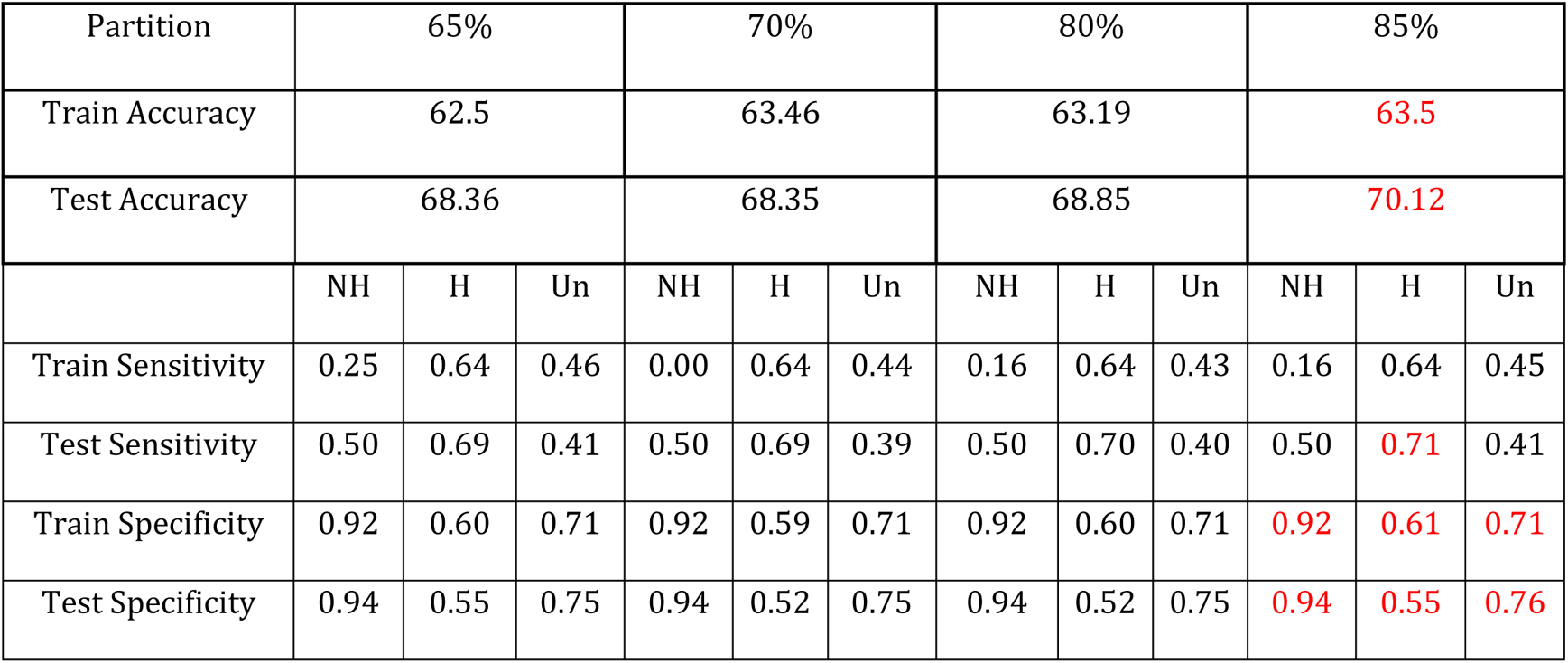
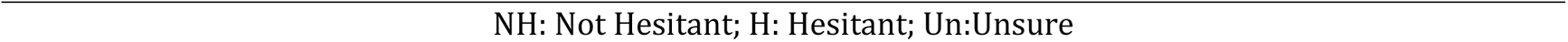
Performance of Prediction Models by train-test partitions.

| Partition | 65% |  |  | 70% |  |  | 80% |  |  | 85% |  |  |
| --- | --- | --- | --- | --- | --- | --- | --- | --- | --- | --- | --- | --- |
| Train Accuracy | 62.5 |  |  | 63.46 |  |  | 63.19 |  |  | 63.5 |  |  |
| Test Accuracy | 68.36 |  |  | 68.35 |  |  | 68.85 |  |  | 70.12 |  |  |
|  | NH | H | Un | NH | H | Un | NH | H | Un | NH | H | Un |
| Train Sensitivity | 0.25 | 0.64 | 0.46 | 0.00 | 0.64 | 0.44 | 0.16 | 0.64 | 0.43 | 0.16 | 0.64 | 0.45 |
| Test Sensitivity | 0.50 | 0.69 | 0.41 | 0.50 | 0.69 | 0.39 | 0.50 | 0.70 | 0.40 | 0.50 | 0.71 | 0.41 |
| Train Specificity | 0.92 | 0.60 | 0.71 | 0.92 | 0.59 | 0.71 | 0.92 | 0.60 | 0.71 | 0.92 | 0.61 | 0.71 |
| Test Specificity | 0.94 | 0.55 | 0.75 | 0.94 | 0.52 | 0.75 | 0.94 | 0.52 | 0.75 | 0.94 | 0.55 | 0.76 |

## Discussion

Several other variable selection methods such as Lasso, Elastic-Net were utilized which did not produce any potential increase in accuracy.

A 2021 study with Robinson et al. concluded that intentions to take the COVID-19 vaccine have been declining across 13 countries. Using a multinomial logistic regression model, we found that gender, geography region, income, marital status, race, worry, education level, hearing ability, and sight were all variables in vaccine hesitancy. Our analysis showed that being male, White, living in the South/Northeast or in a rural area, having low to middle income, and single were more likely to be associated with vaccine hesitancy. Meanwhile, females and Asians were less likely to be associated with vaccine hesitancy. Out study also analyzed vaccine hesitancy in India and China.

Results showed that, similar to the US, the rural population in Indian was more likely to be vaccine hesitant (OR 3.46). In addition, low-income groups in India and China were also more hesitant towards taking the vaccine. The Northern region in China was most likely to be vaccine hesitant. By comparing vaccine hesitancy data from the US to India and China, we see that there are some common factors that contribute to vaccine hesitancy. In all three countries, low income and a rural geographic area are associated increased vaccine hesitancy.

A 2021 questionnaire found that those living in rural areas, having lower incomes, and lower levels of education were more likely you be vaccine hesitant. These results are in concordance with our findings. Khubchandani et al. found that 22% of the respondents to their questionnaire were hesitant to take the vaccine; however, our data shows that 60.9% of the respondents of the HPS survey were hesitant to take the vaccine. Most of the respondents to the HPS survey data used were able-bodied, White, female, having high income, and living in the West or South.

Trogan and Profski posit that overcoming vaccine hesitancy will require a pronged approach. In addition to identifying sociodemographic characteristics that are more likely to be vaccine hesitant, as we have done in this study, the reason behind vaccine hesitancy must also be addressed. Based on results from our study, public health officials and policy maker can target educational and policy interventions to the more hesitant groups to alleviate the reasons behind the vaccine hesitancy and encourage vaccine uptake.

## Conclusion

The ICPSR data gives an overview of the vaccine hesitancy condition on an international level. Also, it informs the health policy and insurance policy makers a guidance on the level of unvaccinated people in the world, and necessary preparations for such a disease.

The cross-sectional survey through the ICPSR database provides vaccine hesitancy on a global scale, comparing different cultures and how these regions and demographics may affect hesitancy. Each country can be analyzed to determine which hesitancy rates are lowest and provide a reasonable explanation. Chi-square tests and exploratory data analysis were conducted on the United States Census Bureau’s Household Pulse Survey (HPS) due to its range of variables surveyed. The HPS data provides a large national sample size, useful when analyzing several variables, while maintaining the survey’s integrity. Using these two data sets, adamant research can be done according to which demographics, region, and mental health related groups produce the highest relative vaccination hesitancy. These findings can be applied to mitigate future pandemics according to vaccine hesitancy rates and globally take steps to place preventative measures for further outbreaks.

**Table 3:**
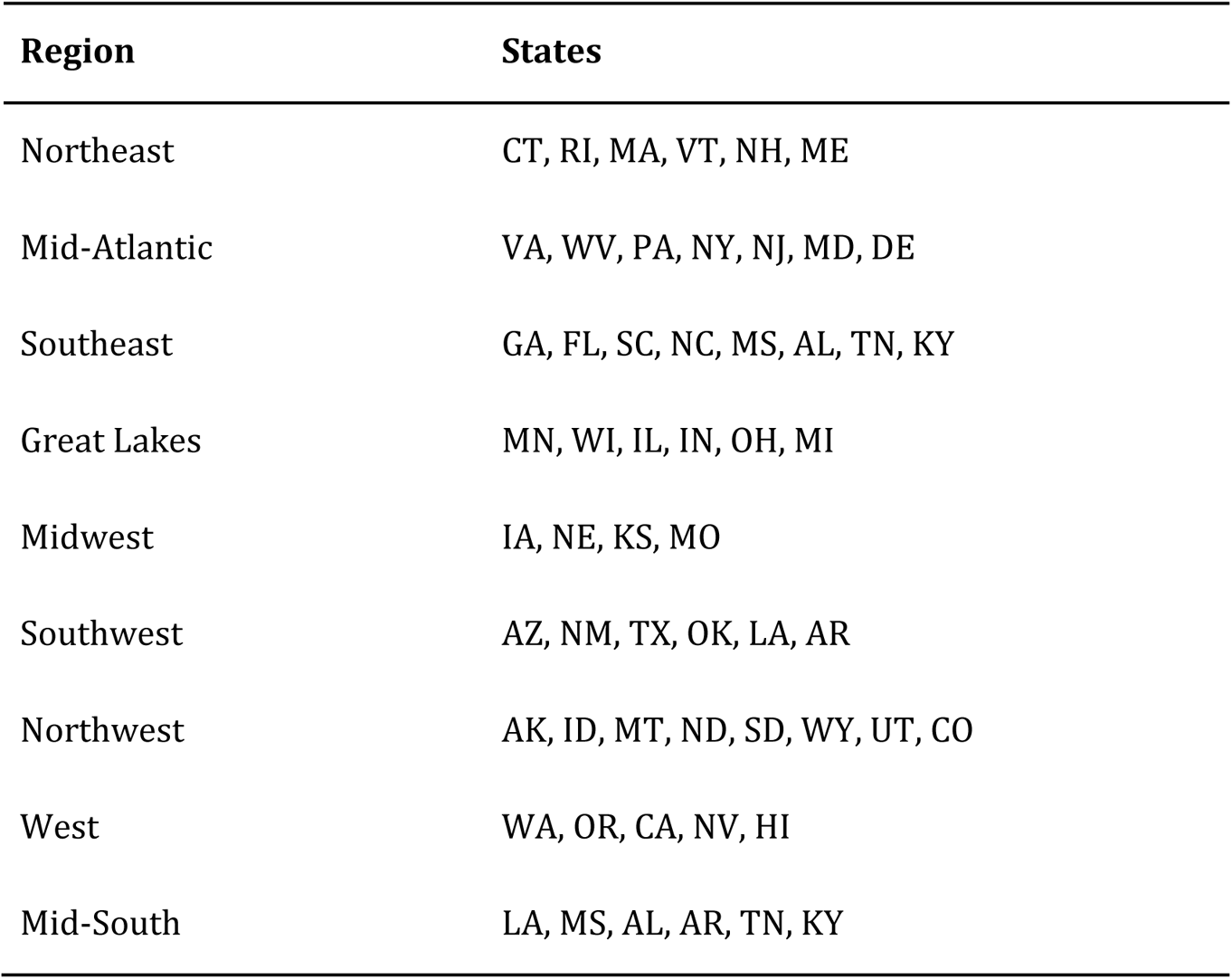
Regions of the US.

## Figures

**Figure 1.**
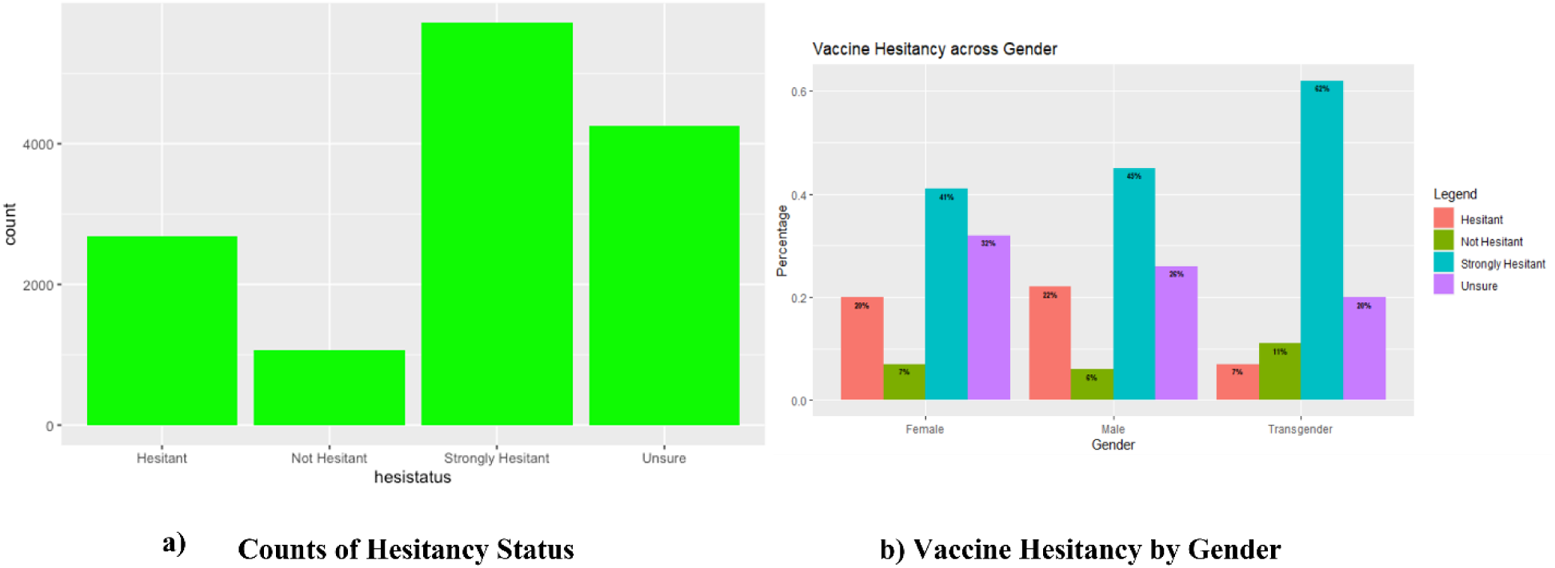

**Figure 2.**
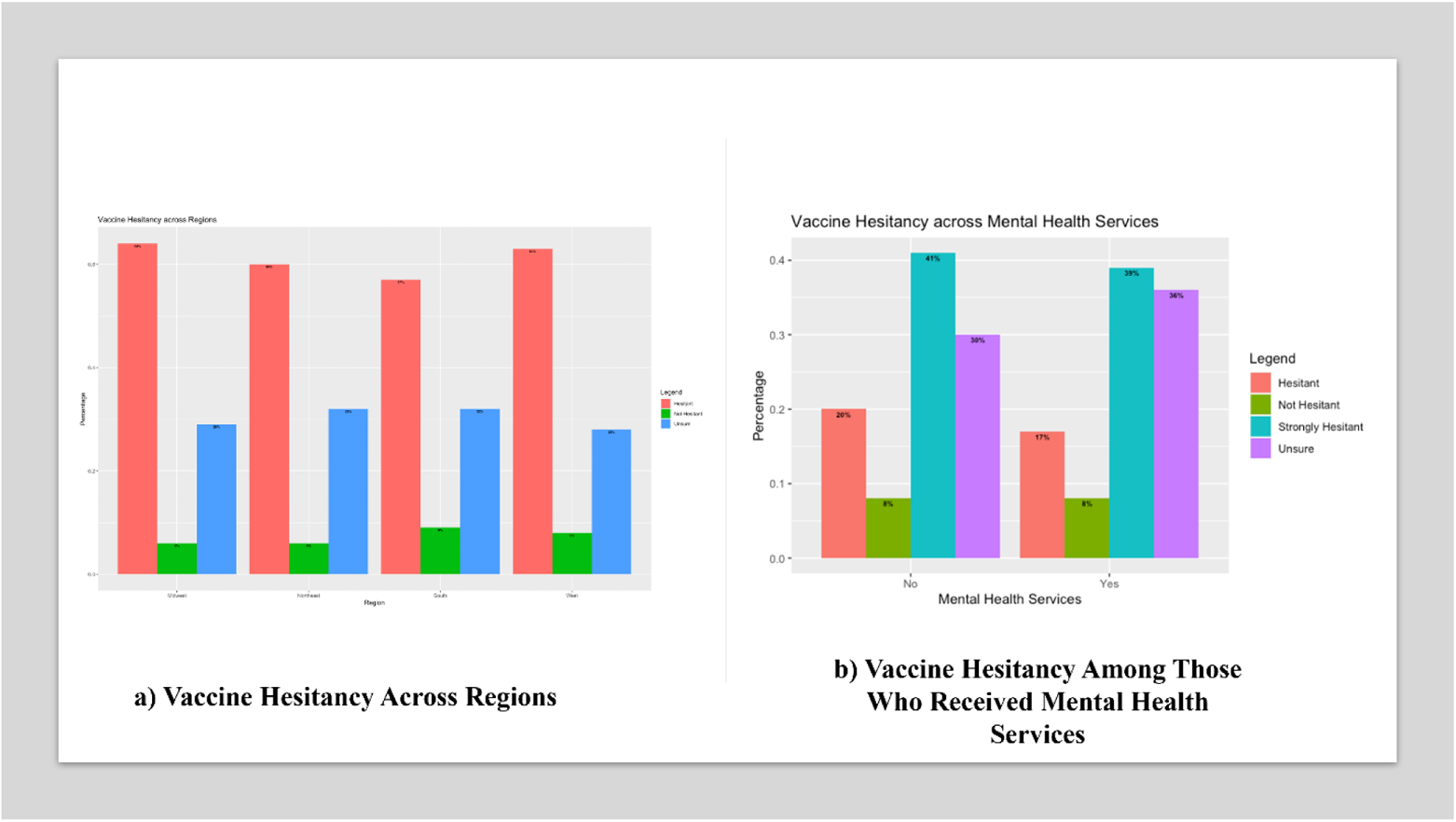

**Figure 3.**
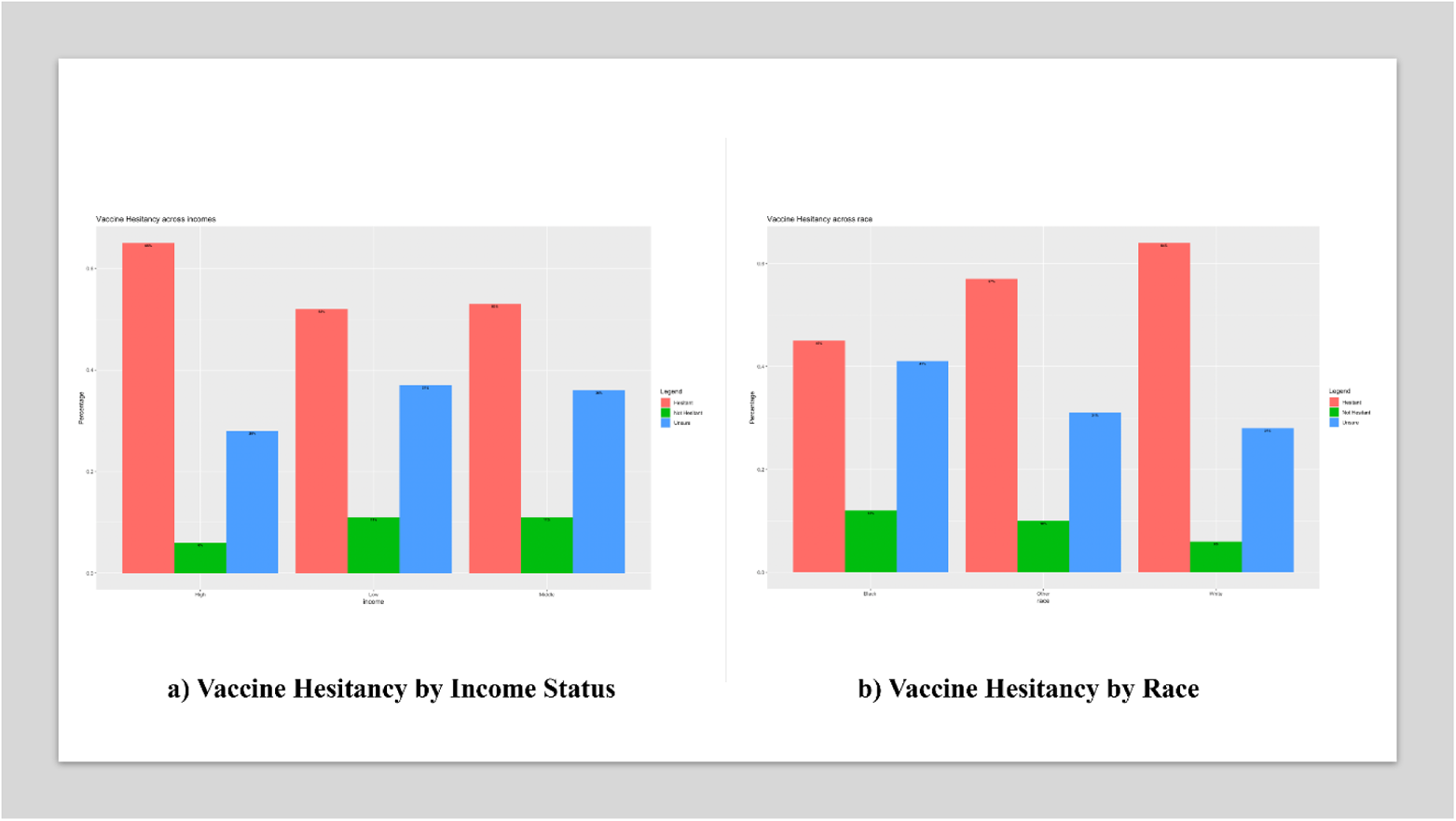

**Figure 4.**
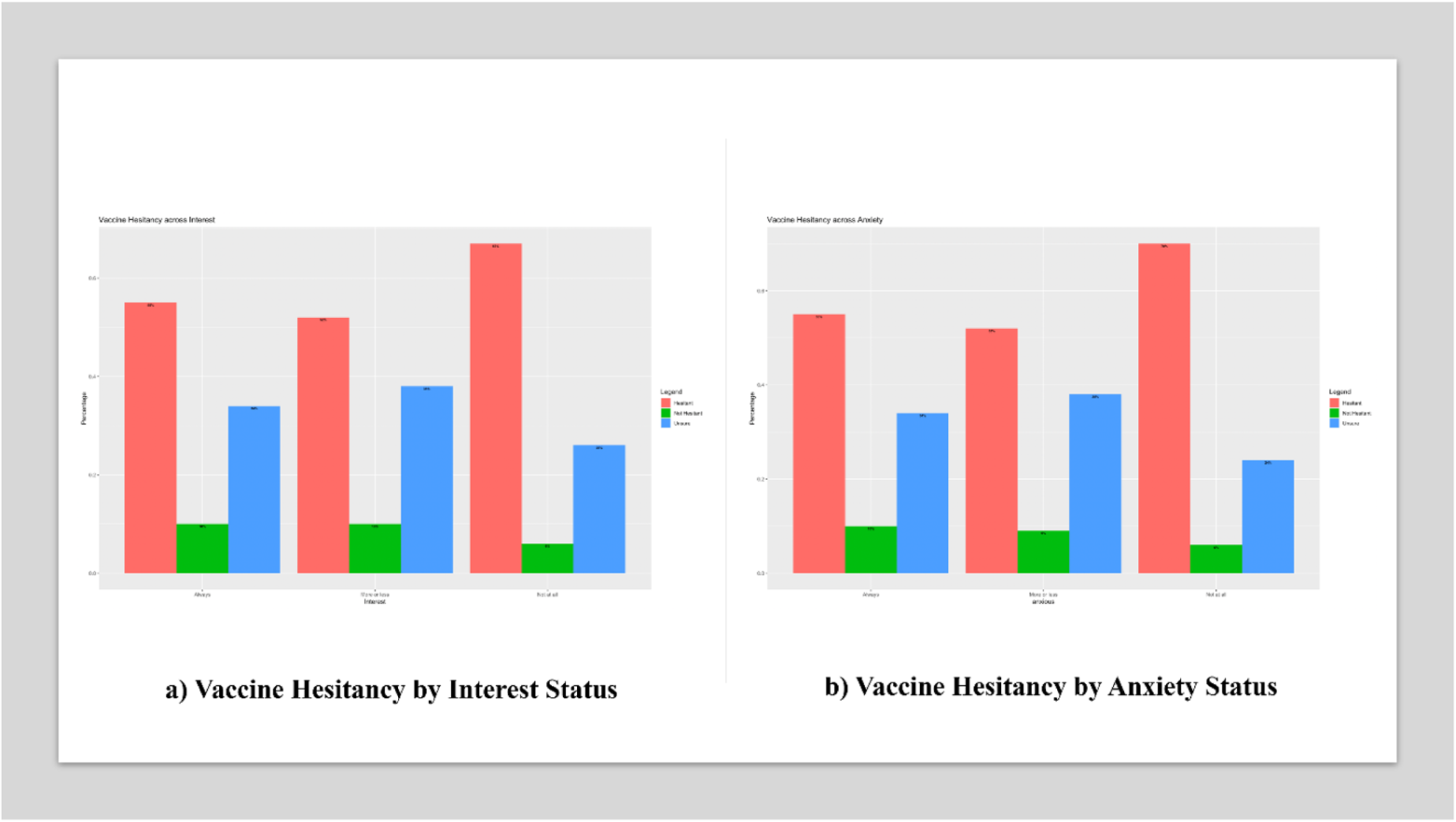

**Figure 5.**
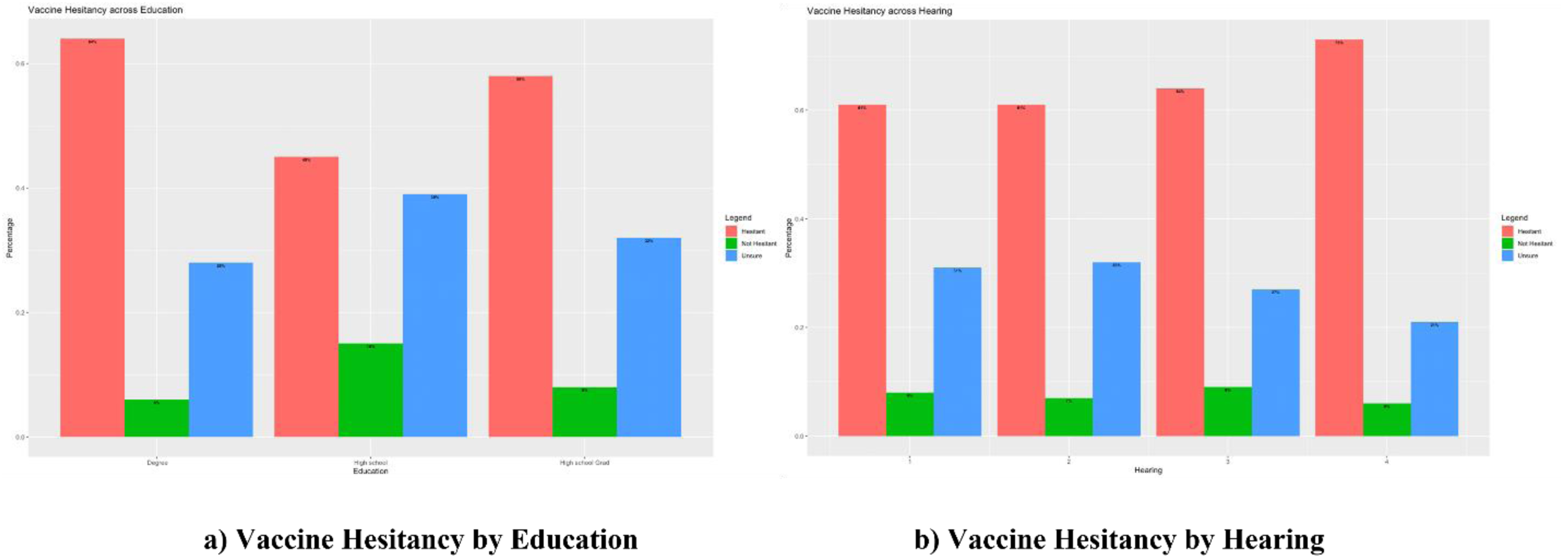

**Figure 6.**
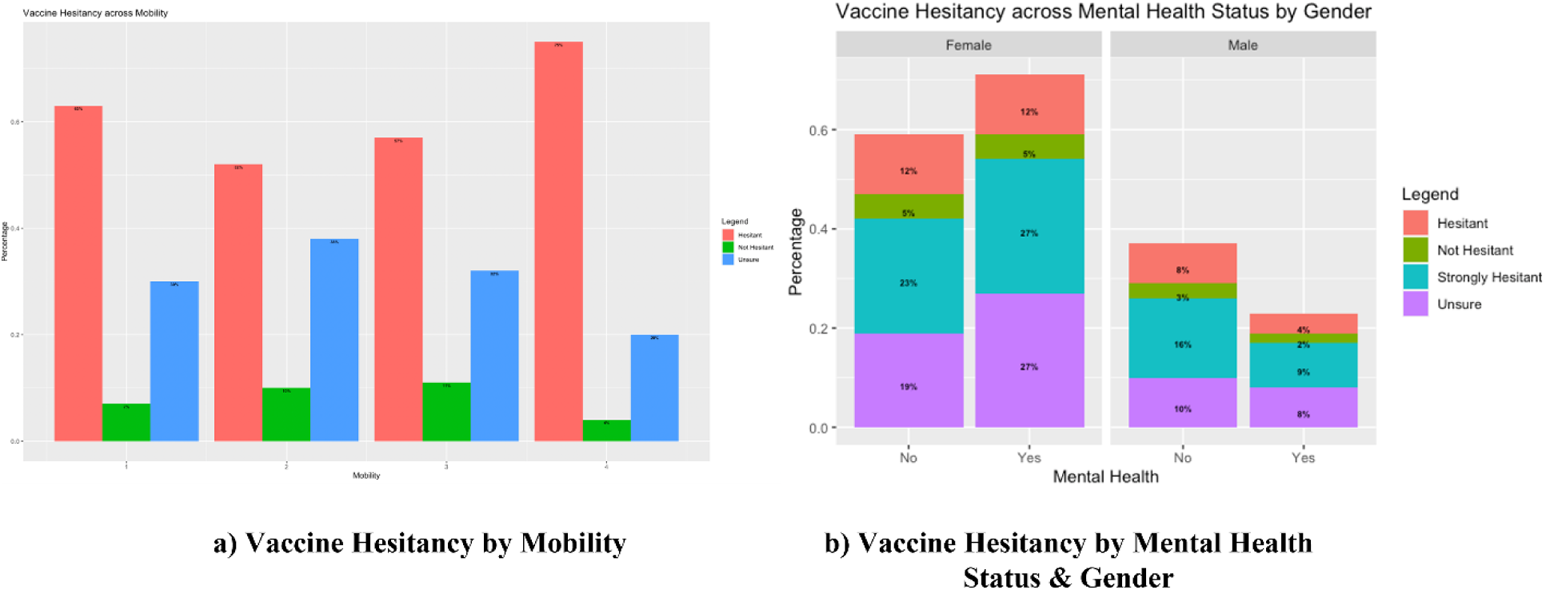

**Figure 7.**
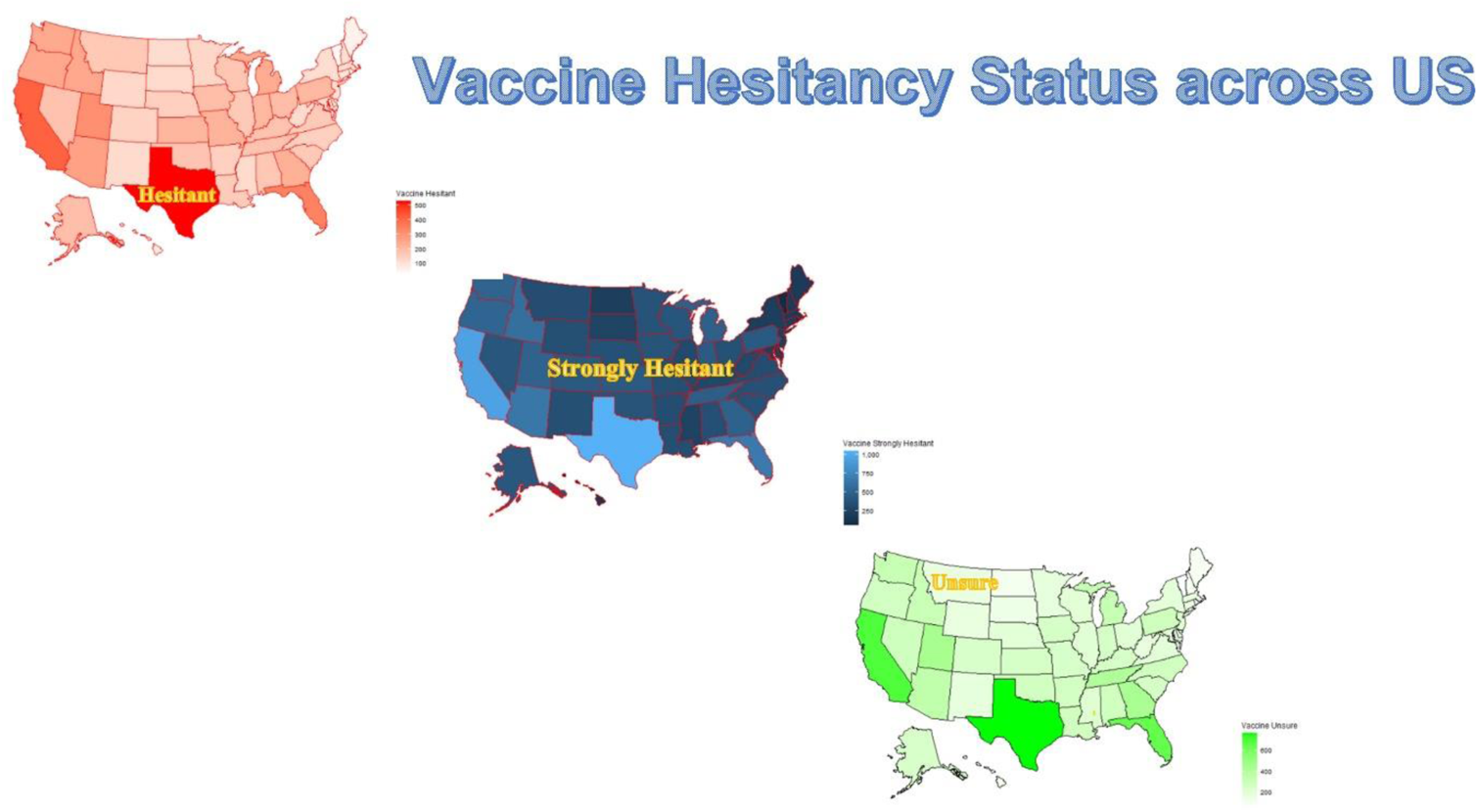

## Data Availability

The first dataset is extracted from ICPSR Covid-19 database (https://doi.org/10.3886/E130422V1) [1]. The second data set is available from https://www.census.gov/data/experimental-data-products/household-pulse-survey.html

https://www.census.gov/data/experimental-data-products/household-pulse-survey.html

## Acknowledgement

The authors would like to thank the reviewers for their valuable comments, and Dr. Seth, and Dr. Rai for their valuable inputs.

